# Midline head position for preterm infants in the first 72 hours of life: A pilot randomised control trial

**DOI:** 10.1101/2025.03.11.25323743

**Authors:** Traci-Anne Goyen, Hannah Skelton, Daphne D’Cruz, Rajesh Maheshwari, Bronwyn Edney, James Marceau, Patricia Viola, Melissa Luig, Dharmesh Shah, Pranav R Jani

## Abstract

**Aim:** Midline head positioning for preterm infants in the first 72 hours of life may prevent intraventricular hemorrhage (IVH). The feasibility of conducting a RCT was explored, namely (1) acceptability of the recruitment and consenting process, (2) practicality of recruitment within 4 hours of life, (3) protocol compliance, and (4) staff satisfaction with the intervention.

**Methods:** An open-label, single center, balanced 1:1 allocation, parallel-group pilot RCT was adopted. Inborn infants <29 weeks admitted to the NICU with no IVH on screening ultrasound and parental consent obtained within 4 hours of birth were randomized to either midline head and supine body position (intervention) or variable position (control) for 72 hours, stratified according to gestation. Measures were recruitment rate, time to complete recruitment, protocol compliance audit, and staff satisfaction survey.

**Results:** Sixty participants were enrolled with recruitment rate of 67%. Recruitment and intervention were commenced by 6 hours. Compliance was 98% for midline head position. Nursing satisfaction was positive in 30/33 (91%). No safety issues were reported for stability, skin integrity, comfort, pain, and head preference.

**Conclusion:** It is feasible and safe to conduct a RCT to examine the neuroprotective effects of positioning the preterm infant in the first 72 hours of life.

**What is already known:**

1. Intraventricular haemorrhage (IVH) is common in preterm infants, mostly occurring within the first 72 hours of life.
2. Head rotation may lead to IVH, and IVH prevention bundles often include midline head position, despite limited evidence.
3. Previous clinical trials have faced challenges and terminated early.

**What this paper adds:**

1. This pilot RCT found midline head positioning is a low-risk intervention and the protocol was feasible and safe to implement.
2. Using explicit consenting, inborn participants were recruited and intervention started within 6 hours after birth.
3. This study informs the design of future comparative effectiveness trials by including outborn infants and waiver of consent, to start the intervention immediately after birth.

## Introduction

Intraventricular haemorrhage (IVH) is common in preterm infants, especially those born <29 weeks gestational age (GA). Across the Australian and New Zealand Neonatal Network, 34.5% of 1,581 infants born <29 weeks had IVH^1^. Severity is associated with poorer neurodevelopmental outcomes in early childhood ^2,3^.

In most cases, IVH occurs within the first 72 hours of life ^4,5^. Fluctuation in cerebral blood flow (CBF) and immature autoregulation contribute to IVH, and subsequently to brain injury^6^. Therefore, reducing fluctuation in CBF is essential to prevent IVH. Head positioning may be associated with IVH ^7,8^. Head rotation (side head turn) may compress the internal jugular vein, leading to venous blood flow obstruction, venous congestion, and fluctuation in CBF. Keeping the head in a midline position with the body may reduce fluctuation in CBF and IVH. Evidence for midline head positioning is emerging, mostly from quality improvement studies^9-11^. Despite the absence of high-quality evidence to support this practice, midline head positioning is often included in IVH prevention bundles. A recent systematic review highlighted the need for a high quality clinical trial on midline head positioning before a change in clinical practice^12^.

A well-designed randomised, controlled trial (RCT) is warranted however enrolling babies born extremely early in a clinical trial soon after birth can be challenging^13,14^. This pilot study explored the feasibility of conducting a future RCT to determine the effects of midline head and supine body positioning for the first 72 hours of life to prevent IVH in preterm infants born <29 weeks GA. The primary objectives of this pilot RCT were to examine the acceptability of the recruitment and consent process to parents, the practicality of recruiting participants within 4 hours of life, compliance with the intervention protocol, and staff satisfaction with the intervention protocol.

From our previous experience, we pragmatically hypothesized 60% of potential participants would be enrolled by 4 hours and at least 80% compliance with the intervention protocol.

## Methods

We conducted an open-label, single centre, balanced 1:1 allocation, parallel-group pilot RCT. Infants were stratified to 2 GA-based groups (Group 1: 23^+0/7^– 25^+6/7^ or Group 2: 26^+0/7^– 28^+6/7^ weeks).

The study was conducted at a tertiary referral hospital delivering perinatal services to approximately 5,000 women annually. Long-term neurodevelopment outcomes were obtained from our follow-up clinic, where this data is routinely collected.

Inborn preterm infants born <29 weeks GA were eligible for inclusion if parental consent was obtained either antenatally or within 4 hours of birth and there was no IVH on a screening cranial ultrasound within 4 hours of birth. We excluded infants if they were outborn or had lethal congenital anomaly, serious congenital cardiac disease, or pre-existing IVH identified on screening ultrasound.

We initially excluded from our study infants with “a need for full cardiopulmonary resuscitation (CPR) at birth” and those with “hypoxic ischemic encephalopathy (HIE)” based on the exclusion criteria of Al-Abdi et al^5^. However, after 2 infants had been enrolled in our study, Kochan et al^15^ reported a significant reduction in severe IVH with head positioning intervention, including infants who had CPR and or HIE. As these infants are at higher risk of fluctuating CBF and therefore developing IVH, we obtained ethical approval to include these infants.

The medical or research staff randomised patients to either the intervention or the control group, by opening a consecutively numbered opaque envelope (a separate set was used for each stratum). The randomisation codes were generated using the permuted blocks method. Multiple births were randomised to the same group.

Mothers at risk of premature delivery admitted to the antenatal ward or the delivery suite were identified by the clinical staff to potentially meet the inclusion criteria. A research team member then introduced the study and obtained parental consent. For non-English speaking families, we used the hospital’s interpreter service. Within 4 hours of birth, infants were screened for study eligibility by trained medical staff performing a cranial ultrasound. When possible, parental consent was obtained antenatally and, when not possible, due to emergency delivery, the consent was obtained within the first 4 hours of birth.

Infants were randomly assigned to either midline head and supine body position (intervention) or variable (control) positioning. A bedside laminated chart reminded staff about the allocated group. The bedside nurse completed a checklist at the end of their shift to document protocol adherence, any problems encountered with the allocated positioning, and reasons for the change if the intervention was discontinued.

The intervention group were positioned supine with the head and the body midline. All intervention infants had the head elevated (incubator bed maximum tilt of 12°) for the first 72 hours of life. The Z-Flo™ (Mölnlycke Health Care, Gothenburg, Sweden) neonatal positioner was moulded, placed under the head and upper shoulders for stable and consistent positioning, including supporting the base of the occiput to prevent forward flexion of the head to avoid airway obstruction (Figure 1). The positioner has a low friction surface, maximum contouring ability for moulding to accommodate tubing, and zero memory, which ensures it does not move once moulded. Every 4 hours, to check skin integrity, infants were briefly turned to side-lying with head and body maintained in the neutral midline using the support of the Z-Flo™ (Figure 1).

**Figure 1:**
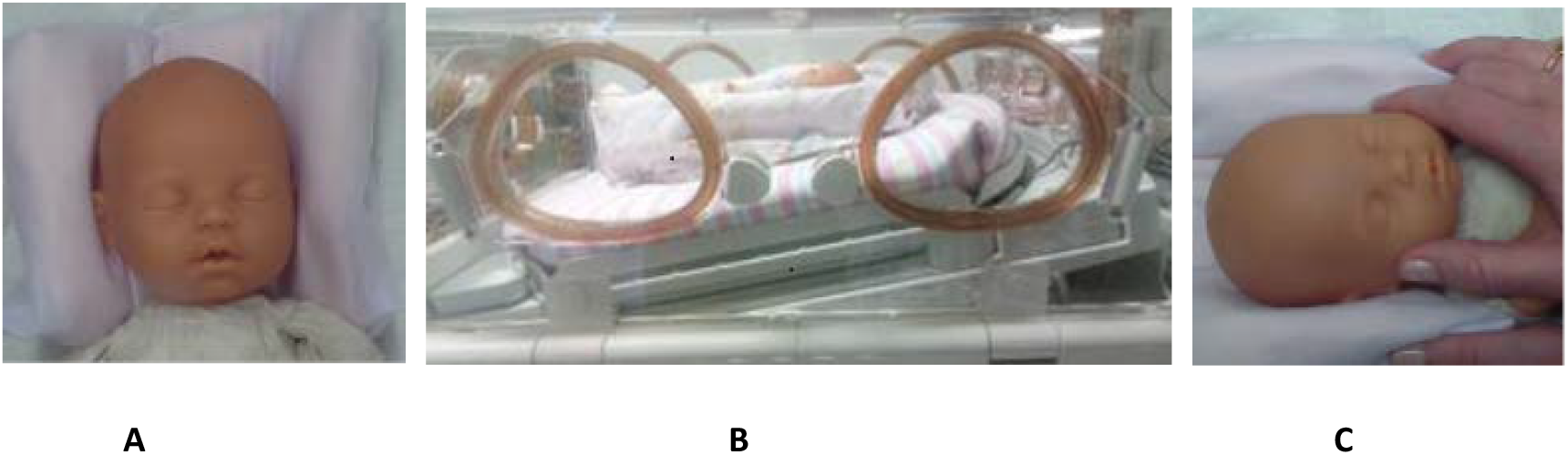
Positioning for intervention group. **A:** Head supported in midline by Z-Flo™ fluidised positioner **B:** Crib tilted at a 12° angle **C:** For caregiving, head and positioner is turned together to avoid head rotation

Infants in the variable position (control) group had no specific instructions for head and body position or for crib tilt. Our unit did not have a standardised head and body positioning practice and no attempt was made to standardise the early head positioning practices during the study period.

A. The primary objectives/outcomes of the feasibility study were to examine the:
  1. Acceptability of the recruitment/consenting process to parents, using the recruitment rate and parent survey,
  2. Practicality of recruitment within 4 hours of life,
  3. Compliance with the intervention protocol using an audit, and
  4. Staff satisfaction with the intervention protocol using a staff survey.
B. The secondary objective/outcome was:
  5. Rates of IVH in infants born <29 weeks gestation on cranial ultrasound between the two groups
C. Tertiary objectives/outcomes:
  6. Rates of abnormality on early marker of cerebral palsy using General Movements Assessment^16^,
  7. Early motor ability on Test of Infant Motor Performance^17^,
  8. Neurodevelopmental outcome (cognitive and motor scores) with Bayleys Scales^18^,
  9. Rates of cerebral palsy diagnosis by a paediatrician
D. Ancillary and safety objectives examined were:
  (i) Physiological stability
  (ii) Skin integrity^19^
  (iii) Pain^20^
  (iv) Comfort^21^
  (v) Early head preference^22^

The objectives and outcome measures are detailed in Table 1.

**Table 1:** Summary of objectives and outcome measures for the RCT.

| **Primary Objectives** | **Outcome Measure** |
| --- | --- |
| Acceptability of recruiting and consent process to parents | The number of infants consented/recruited were compared to those who were eligible but could not be recruited or did not consent, and the parent survey |
| Practicality of recruiting participants within 4 hours of life | Time to complete recruitment was recorded by bedside nurse/research nurse |
| Compliance with intervention protocol | Documentation of study compliance/alterations (eg. bed flattening for clinical procedures) completed at the end of the shift by the bedside nurse and compliance audits conducted by research nurses twice in the 72-hour period |
| Staff satisfaction with the study protocol | Explored using a staff survey of NICU nurses which was completed after the recruitment period |
| **Secondary Objective** |  |
| Prevention of IVH (any grade) | Proportion with IVH (any grade) on cranial ultrasound as determined by independent radiologist by 5 days of life |
| **Tertiary Objectives** |  |
| Reduced abnormality on early marker of cerebral palsy | Abnormality on General Movements Assessment at 32, 36 weeks and 3 months corrected age |
| Improved early motor ability | Abnormality on Test of Infant Motor Impairment at 36 weeks and 3 months corrected |
| Improved neurodevelopmental outcome (cognitive and motor scores) at 2 years | Abnormality on Bayley Scales of Infant and Toddler Development 4<sup>th</sup> Ed<sup>18</sup> (cognition and motor scores) at 2 years corrected age |
| Reduced diagnosis of cerebral palsy at 2 years | Proportion clinically diagnosed with cerebral palsy at 2 years |
| **Ancillary Objectives – Safety** |  |
| Physiological stability of infants | Episodes of accidental extubation, apnea, apnea requiring intubation or IPPV hypotension requiring inotropes from medical records. |
| Skin integrity from positioning over 72 hours | Neonatal Skin Condition Score<sup>19</sup> was measured at each caregiving episode. The erythema rating was analyzed to provide an indication of pressure on skin. Erythema is scored on a 9-point, 0 – 4 scale with 0.5-grade increments. |
| Pain | Neonatal Pain Assessment Tool (PAT)<sup>20</sup> completed at each caregiving episode. This is an observational scale designed to assess neonatal pain from 24-weeks gestation to term age, and up to 6-months old. Scores from 0 - 4 indicate comfort, and scores above 4 indicate pain/discomfort. |
| Comfort of positioning | COMFORT-NEO<sup>21</sup> was completed at 24, 48 and 72 hours by either the research nurse or medical registrar. It is an observation tool with 7 categories: alertness, calmness/agitation, respiratory response (mechanically ventilated only), crying (spontaneously breathing infants only), body movement, facial tension, and (body) muscle tone. Scoring in each category ranges from 1 – 5, with a total cut-off score of 14 indicating pain/distress. |
| Early head preference | Any early head preference was recorded using the Head Turn Preference Scale<sup>22</sup> which was administered between 36 – 38 weeks gestation. Scores range from 0 - 10, which are categorized as no (score of 0), minimal (scores of 1 – 3), and moderate/severe head turn preference (scores 4 - 10). |

IPPV = Intermittent positive pressure ventilation

The independent assessors for secondary and tertiary outcome measures were blinded to group allocation at all measurement points. All assessments were carried out by blind assessors after the intervention had been completed.

## Data Analysis

A sample size of 60 infants with a 1:1 allocation ratio was considered a representative sample to determine the feasibility outcomes and was consistent with recommendations for feasibility studies^23^. This study was not powered to answer the secondary or tertiary objectives and these are not reported.

Descriptive statistics were used to report feasibility outcomes (ie. primary objectives and ancillary measures). Data were summarized as frequencies and proportions or as free text. All analyses were based on the intention-to-treat principle and performed using SPSS software version 21.0^24^. Demographic and clinical characteristics were summarised by descriptive statistics. Normally distributed continuous variables were summarised as mean and standard deviation and, where not normally distributed, were summarised as median and interquartile range (IQR). Categorical variables were summarised as proportion and count.

This study was approved by the local Ethics Committee (HREC/18/WMEAD/129).

## Results

### Participant Flow and Recruitment

Recruitment commenced on 13 February 2019 and ceased on 29 July 2021, including a 9-month mandatory suspension for the COVID-19 pandemic.

Figure 2 shows the consolidated standards of reporting trials (CONSORT) diagram for patient flow. There were 105 infants who delivered at <29 weeks gestation and were admitted to the NICU. Of these, 30 infants met the pre-specified exclusion criteria. Consent was not obtained for a further 7 due to an emergency delivery, and 8 were not enrolled for other reasons such as complex social histories (n=3), critical condition of mother/baby (n=3), unknown or incorrect gestation (n=2).

**Figure 2:**
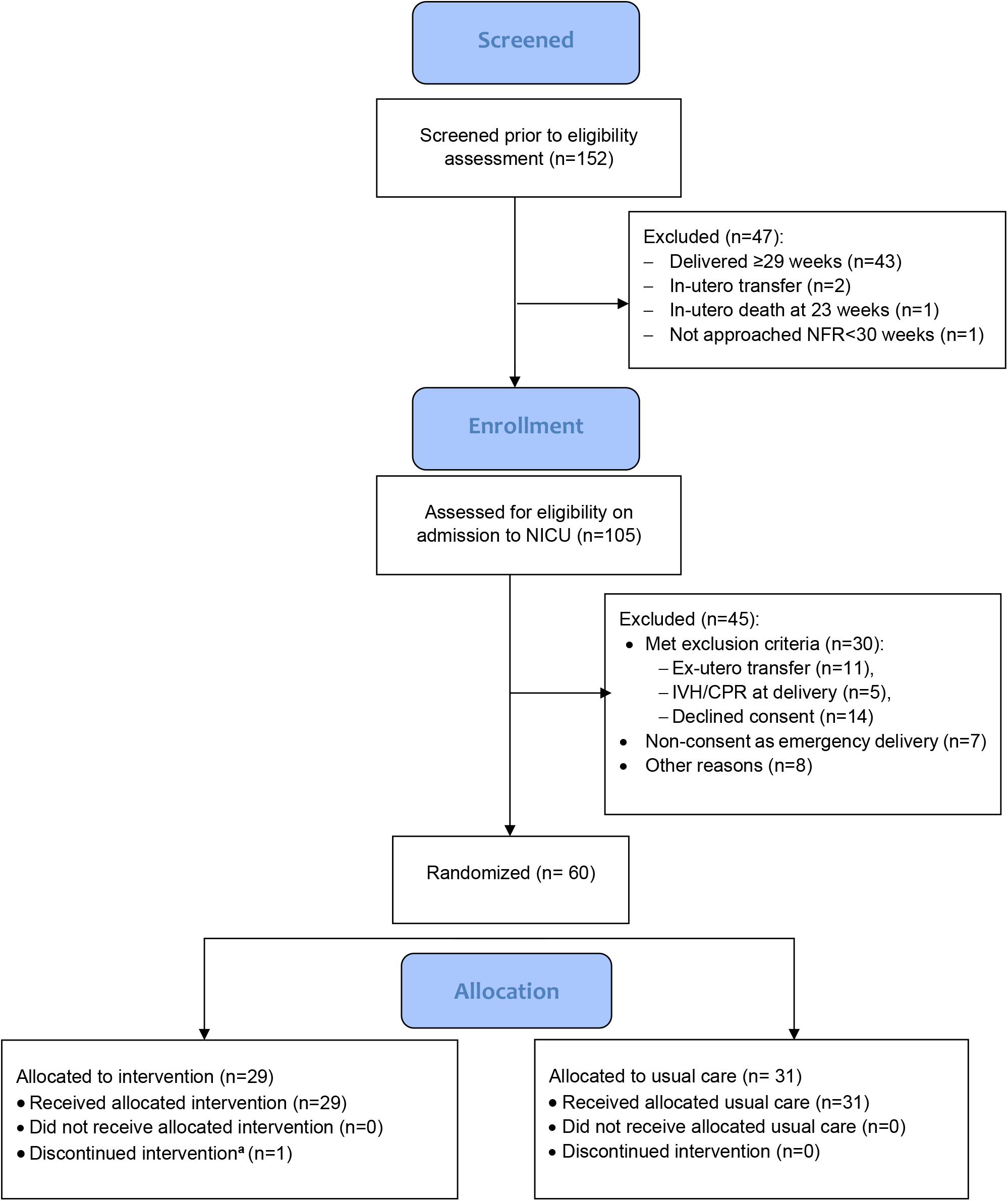
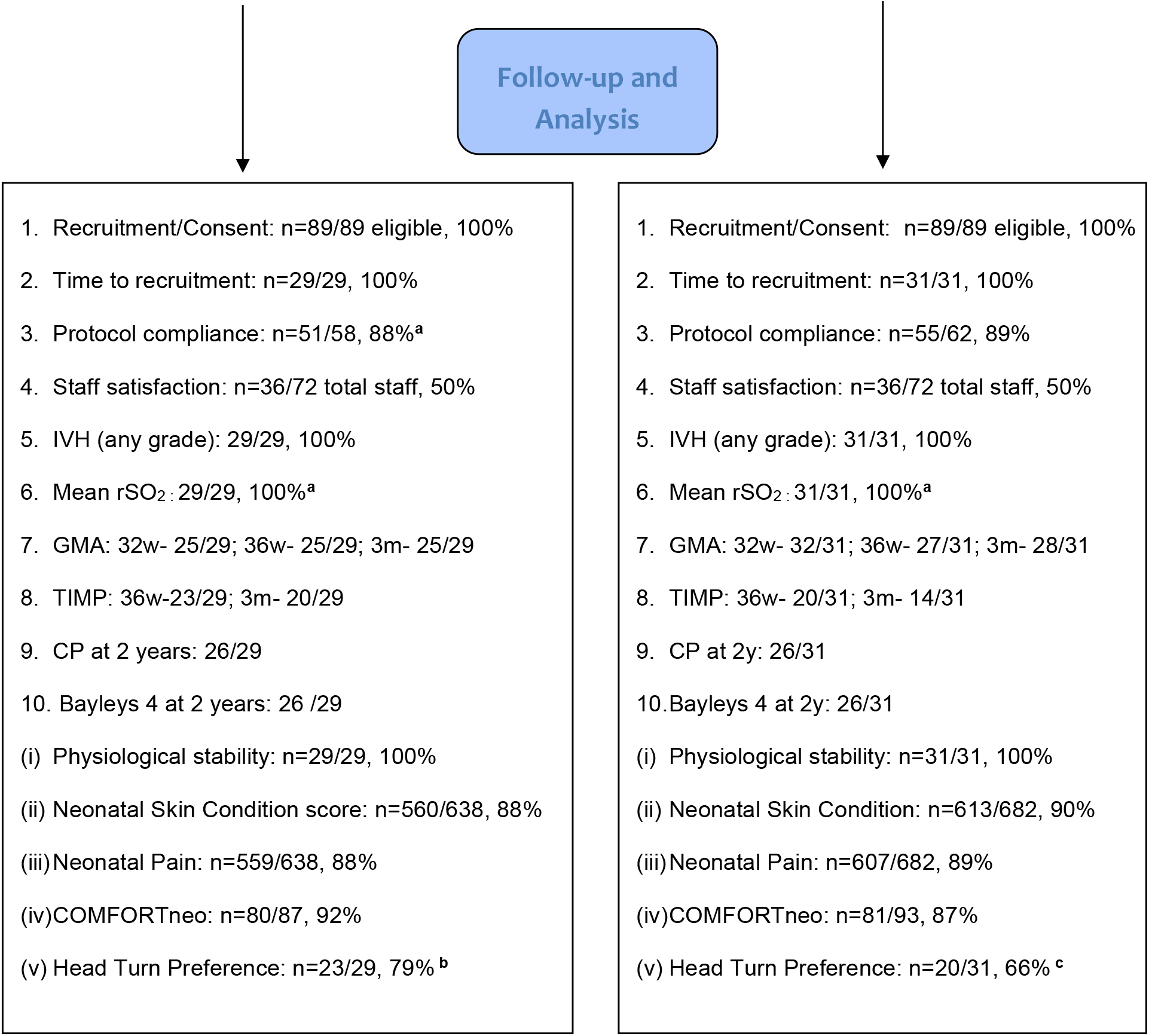
Flow of study participants. ^**a**^ One baby received only 48 hours of intervention due to clinical deterioration ^**b**^ n=5 transferred prior to 36 weeks ^**c**^ n=10 transferred prior to 36 weeks IVH=intraventricular haemorrhage, rSO2=cerebral perfusion; w=weeks; m=months; GMA=General Movements Assessment; TIMP=Test of Infant Motor Performance; CP=cerebral palsy

The remaining 60 eligible infants were enrolled [intervention group (n=29) and control group (n=31)] and included in the final analysis. Although we had planned to have 30 infants in each group, a set of triplets were allocated to the same treatment group (control group), which led to the imbalance in groups. Baseline characteristics for both groups were similar (Table 2).

**Table 2:** Baseline characteristics of participants.

| Variables | Intervention (n = 29) | Control Group (n = 31) |
| --- | --- | --- |
| Gestation (weeks) | 27 (26,28) | 27 (25.75,27.25) |
| Birthweight (grams) | 990 (826.25,1178.2) | 901.5 (830.5,1030) |
| Antepartum haemorrhage | 8 (28.6%) | 9 (28.1%) |
| Antenatal steroids Complete n(%) | 18 (64.3%) | 25 (78.1%) |
| Multiple birth n(%) | 6 (21.4%) | 10 (31.3%) |
| Chorioamnionitis | 5 (17.9%) | 5 (15.6%) |
| Caesarean section | 20 (71.4%) | 19 (59.4%) |
| Males n (%) | 15 (53.6) | 18 (56.3) |
| Intrauterine growth restriction (<10<sup>th</sup>%) | 2 (7.1%) | 3 (9.3%) |
| 5-minute Apgar <6 | 4 (14.2%) | 5 (15.6%) |
| Retinopathy of prematurity (stage≥3) | 4 (14.3%) | 2 (6.3%) |
| Died prior to discharge | 1 (3.6%) | 0 (0%) |

All participants received the intended treatment for 72 hours, except for 1 infant in the intervention group. The attending medical team ceased the intervention after 48 hours due to clinical deterioration and decision for redirection of care. This deterioration was not considered to be a result of the intervention.

### Primary Outcomes

Primary outcomes and ancillary analyses are summarized in Table 3.

**Table 3:** Summary of primary outcomes and ancillary analyses for study groups.

|  | FEASIBILITY OBJECTIVE | RESULTS |  |  |
| --- | --- | --- | --- | --- |
| 1. | **Recruitment/Consent process:**<br>Number consented / total eligible | 60 / 89 (67%) |  |  |
| 2. | **Recruitment within 4 hours:**<br>Mean time to recruit,<br>hours±SD (range)<br>Number recruited within 4 hours<br>Number recruited within 6 hours | <u>Intervention</u><br>3.54±0.87<br>(1.78-5.67)<br>22/29 (76%)<br>29/29 (100%) | <u>Control</u><br>3.34±1.0<br>(1.18-5.43)<br>20/31 (64%)<br>31/31 (100%) | <u>p-value</u><br>0.41<br>0.49 |
| 3. | **Protocol compliance:**<br>Positioned on pillow<br>Head midline and shoulders rotated forward<br>Crib tilted (maximum tilt)<br>NIRS in situ | <u>Intervention</u><br>(n = 51 audits)<br>51 (100%)<br>50 (98%)<br>43 (84%)<br>51 (100%) | <u>Control</u><br>(n = 55 audits)<br>N/A<br>N/A<br>N/A<br>54 (98%) |  |
| 4. | **Staff satisfaction with intervention:** | Total 36 surveys completed |  |  |
|  | Education received prior to study | Not enough = 8<br>Just right = 28 |  |  |
|  | Did study packs make it easier for study set up? | No = 1<br>Yes = 32<br>Unknown = 3 |  |  |
|  | Documentation during study | Acceptable number and easy to complete = 21<br>Too many forms but easy to complete = 12<br>Too many forms and hard to complete = 1<br>Unknown = 2 |  |  |
|  | Able to support head in midline with Z-Flo positioner | Yes = 30<br>No = 2<br>Only controls and did not use = 3<br>Unknown = 2 |  |  |
| i. | **Physiological stability:**<br>Number of accidental extubation episodes<br>Mean episodes of apnoea (IQR)<br>Episodes of apnoea requiring intubation<br>Mean episodes of apnoea requiring IPPV (IQR)<br>Episodes of hypotension requiring inotropes | <u>Intervention</u><br>0<br>3 (0 - 5.5)<br>3<br>0 (0 - 1)<br>0 | <u>Control</u><br>1<br>0 (0 - 1)<br>1<br>0 (0 - 1)<br>1 | <u>p-value**</u> |
| ii. | **Neonatal Skin Condition Score**<br>Mean (±SD) | <u>Intervention</u><br>(n = 560)<br>0.54 (±0.69) | <u>Control</u><br>(n = 613)<br>0.60 (±0.71) | <u>p-value</u><br>0.14 |
| iii. | **Neonatal Pain Assessment Tool** | <u>Intervention</u><br>(n=559 scores) | <u>Control</u><br>(n=607 scores) | <u>p-value</u> |
| | Mean (±SD)<br>0-4 (comfort)<br>>4 (Discomfort/Pain) | 3.13 (±2.63)<br>408 (73%)<br>151 (37%) | 3.06 (±2.08)<br>456 (75%)<br>151 (25%) | 0.61 |
| iv. | **COMFORTneo**<br>Mean (±SD)<br><14 (no pain/stress)<br>>14 (moderate-severe pain/stress) | <u>Intervention</u><br>(n = 80 scores)<br>12.71 (±17.08)<br>54 (67%)<br>26 (32%) | <u>Control</u><br>(n = 81 scores)<br>11.49 (±3.83)<br>59 (73%)<br>22 (27%) | <u>p-value</u><br>0.53 |
| v. | **Head Turn Preference Scale:**<br>No/Minimal<br>Moderate/Significant | <u>Intervention</u><br>(n = 23 scores)<br>18 (78%)<br>5 (21%) | <u>Control</u><br>(n = 20 scores)<br>17 (85%)<br>3 (15%) | <u>p-value</u><br>0.7 |

\*\*p-value not reported due to small event rates; SD=standard deviation; NIRS=Near-Infrared
Spectroscopy: IQR=interquartile range; IPPV= Intermittent positive pressure ventilation

1. Recruitment/consenting: The successful recruitment rate was 67% (60/89).
2. Time for recruitment: The mean time to complete the screening ultrasound was 3.54 hours for the intervention group and 3.34 hours for the control group. The intervention was commenced in 76% infants within 4 hours and in 100% by 6 hours of life.
3. Protocol compliance: All intervention infants were positioned supine, on the Z-Flo positioner, with their head in midline. One infant, on one occasion, had the head positioned slightly from midline. Maximum tilt for the crib was recorded for 84%. One in the control group had kangaroo care for one hour during the first 72 hours.
4. Staff satisfaction: From 36 completed surveys (50% response rate), most staff (78%) indicated they were satisfied with the education, study packs made set up easy (89%) and forms were easy to complete (92%). Of those nursing the intervention participants, most did not have problems using the Z-Flo positioner (91%), commenting that the positioner was easy to mould and the diagrams/photos were easy to follow. One nurse had difficulty with the infant sliding down the positioner.

### Ancillary Analyses: Intervention Safety

i. Physiological stability: Mean episodes of apnoea was significantly lower for the control group (p=0.03), although apnoea episodes requiring intubation or intermittent positive pressure ventilation were not significantly different between groups (p=0.25 and p=0.74, respectively).
ii. Skin integrity: The mean total score was similar for both groups (intervention=0.54, control=0.60, p=0.14).
iii. Pain: Mean pain score was similar between groups (intervention=3.13 vs control=3.06 indicating comfort with no pain, p=0.61) mostly scoring in the comfortable range (73% intervention and 75% control).
iv. Comfort: Mean scores were within the comfortable range (intervention=12.71 vs control= 11.49, p=0.53). Most scores were within the comfortable range (intervention=54/80, 67%; control=9/81, 73%).
v. Early head preference: No to minimal head turn preference was recorded for 78% of the intervention group and 85% of controls (p=0.7).

## Discussion

This feasibility study demonstrated acceptance of the recruitment and consent process by parents (67% recruitment rate), and feasibility of recruiting over 60% of participants within 4 hours of life. There was very high rate of compliance to the protocol, aside from some variability of crib tilt, and there was high staff satisfaction with the intervention. However, it took up to 6 hours for all participants to have a baseline ultrasound, obtain consent and start intervention. While the staff-satisfaction survey response rate was lower than anticipated, the majority indicated that the protocol was acceptable. The RCT was feasible without an increase in adverse events, such as pain, discomfort, and skin injury, or a change in other safety measures. Previous studies midline head positioning have not reported on safety measures^15, 25-26^. As the intervention is frequently used in clinical practice, our results provide additional supporting evidence that midline head and supine body position should be considered a low-risk intervention.

The recruitment rate for this study was reasonable. However, the four-hour time frame for prospective consenting, particularly when obtaining consent in the birthing unit or postnatally, may be too short. Difficulties with a prospective informed consent process for neonatal clinical trials have been reported previously, notably for emergency deliveries, and may lead to discontinuation, underpowered studies or a selection bias limiting the generalisability of outcomes^13, 27^. A similar trial investigating midline head positioning was stopped early due to poor recruitment^26^ and another RCT closed due to slow recruitment^25^. Our recruitment rate of 67% was higher than the comparable Kochan et al^15^ study, which achieved a rate of 59% and missed 113 participants, as prior consent was unavailable. In our study, we had similar challenges for enrolling infants following an emergency delivery despite having a dedicated team approach to capture consenting over the 24-hour period. Ex-utero transfers were excluded and may also have benefitted from this intervention.

Intervention for all infants could only be started by 6 hours. Time to complete a screening ultrasound, particularly for emergency deliveries, contributed to the delay and may increase IVH risk due to further unsupported handling. Timeframe for screening ultrasounds may be too difficult in many NICUs and likely decrease the recruitment rate.

The protocol was successfully implemented in this busy NICU setting, although there was no provision to accommodate kangaroo care and some variation with the tilt of the crib. Kochan et al^15^ also reported numerous protocol violations relating to flattening the bed for procedures. All are potential barriers for recruitment.

We recommend incorporating the following changes when designing future trials to definitively answer the research question: Firstly, inclusion of outborn infants and those with emergency deliveries are vulnerable and potentially benefit from inclusion in a definitive trial. Secondly, an exploration of an alternate consent process such as waiver of consent with a follow-up opportunity for obtaining informed consent for ensuring inclusion of these infants should be explored. Attaining a reasonable-to-high recruitment rate may be difficult for centres when there is no dedicated research team frequently screening antenatal wards and high-risk pregnancies. A four-hour timeframe is considered too short for the processes of obtaining consent and providing a screening ultrasound. In addition, ultrasound screening may not be available universally. Therefore, removing screening ultrasounds may reduce the barrier for starting the intervention immediately after birth and ensure wider inclusion for middle- and low-income countries. This is necessary as approximately 50% of IVHs occur within 6 hours of life^29^. Eliminating bed tilt and accommodating alternate midline positioning for kangaroo care will improve protocol compliance across sites.

This was a single-site unblinded study. Bias was minimised by ensuring education and a 24-hour compliance check during the delivery of the intervention. A significant limitation of this study was the pause for COVID-19 and may have had an impact on the recruitment rate.

## Conclusion

It is feasible and safe to undertake a RCT on midline head and supine body position to prevent IVH for the first 72 hours of life in preterm infants <29 weeks GA. While it is possible to achieve an acceptable recruitment rate, an alternate consenting process, such as a waiver of consent, should be explored for future comparative effectiveness trials. This would maximise the inclusion of all participants, especially outborn infants or those born in an emergency^30^, and improve the generalisability of results. This pilot RCT provides strong feasibility data for designing a future multisite trial.

## Data availability statement

The datasets generated during and/or analysed during the current study are not publicly available due to original ethics committee specifications but are available from the corresponding author on reasonable request and with ethics compliance.

## Ethics Approval Statement

The PIN study received ethical approval from the Western Sydney Local Health District Human Research Ethics Committee (HREC/18/WMEAD/129).

## Patient Consent Statement

Written, informed consent was obtained from parents of all participating infants in this study.

## Clinical trial registration

The PIN study was registered with the Australian and New Zealand Clinical Trials Registry (ACTRN12619000276156).

## Acknowledgement Statement

We would like to acknowledge the contribution of the infants and their families who participated in this study, and nursing staff who provided exceptional care for the babies.

